# Spatial Distribution and Multilevel Determinants of Unvaccinated Children in Somaliland: An Analysis of the 2020 SDHS. Spatial and Multilevel Analysis

**DOI:** 10.1101/2025.09.16.25335900

**Authors:** Hana Mahdi Dahir, Ayan Husein Korse, Saralees Nadarajah

## Abstract

**Background:** Unvaccinated children, often termed ‘zero-dose,’ represent a critical global public health challenge and a stark indicator of health system inequities, particularly in low– and middle-income countries. Somaliland faces an exceptionally severe crisis, with an alarmingly high prevalence of zero-dose children. This study aimed to comprehensively assess the spatial distribution and identify the individual and community-level determinants of unvaccinated children among those under five years in Somaliland, utilizing data from the 2020 Somaliland Demographic and Health Survey (SDHS).

**Methods:** This cross-sectional study analyzed data from 3,255 children under five from the 2020 SLHDS. Multilevel logistic regression analysis was employed to identify determinants of zero-dose status, comparing models using AIC, BIC, and Log Likelihood, and assessing community-level variation with the Intraclass Correlation Coefficient (ICC). Geospatial analyses, including descriptive mapping, Global Moran’s I, Local Moran’s I (LISA), and Getis-Ord Gi* statistics, were used to explore the spatial distribution and clustering of unvaccinated children.

**Results:** The prevalence of unvaccinated children in Somaliland was found to be extremely high at 79.88%. The final multilevel model (Model III) indicated that significant individual-level factors associated with lower odds of being zero-dose included maternal secondary education (AOR=0.409), not currently breastfeeding (AOR=0.565), delivery in a health facility (AOR=0.642), belonging to middle (AOR=0.602) or rich (AOR=0.499) wealth quintiles, and attending any ANC visits (AORs 0.349 for <4 visits; 0.253 for ≥4 visits). Spatial analysis revealed marked regional disparities, with Awdal (88.66%) and Sool (84.71%) exhibiting the highest zero-dose prevalence, while Sahil (71.43%) and Togdheer (75.33%) had the lowest rates. Global Moran’s I indicated overall negative spatial autocorrelation, suggesting a pattern of dissimilarity. Local Moran’s I identified clusters of lower zero-dose rates in Sahil, Maroodi Jeex, and Togdheer, while Awdal and Sool emerged as high-rate outliers. The initial ICC of 24.8% (Model 0) reduced to 7.5% (Model III), confirming substantial community-level influence on zero-dose status.

**Conclusion:** Zero-dose vaccination status is alarmingly prevalent and exhibits significant spatial heterogeneity in Somaliland. Key determinants include maternal education, healthcare utilization (ANC visits, facility delivery), and household wealth. The geospatial patterns highlight specific high-risk regions and areas with clustering of lower rates, underscoring the need for geographically targeted and context-specific interventions to reach unreached children and improve immunization equity.

## INTRODUCTION

Childhood immunization is a cornerstone of global public health, widely recognized as one of the most successful and cost-effective interventions for preventing mortality and morbidity from a host of infectious diseases[1][2][3]. Zero-dose children, as defined by UNICEF (2025), are those who have missed out on all routine immunizations, signifying profound inequities in access to essential health services. [4]. Globally, approximately 6.3 million children under five die each year, with Sub-Saharan Africa disproportionately affected, accounting for about half of the 1.5 million global vaccine-preventable deaths annually [5][6]. While global immunization efforts have impressively saved an estimated 154 million lives over the past 50 years and significantly reduced infant deaths by over 50% in the African Region, monumental challenges remain. During the pandemic years, an estimated 67 million children missed one or more essential vaccines, and coverage for critical vaccines like measles is still below outbreak prevention thresholds, highlighting ongoing vulnerabilities, especially in resource-limited settings[7]

Globally, the challenge of reaching every child with essential immunizations persists, with an estimated 14.5 million children under one year of age not receiving any basic routine vaccine doses in 2023, categorizing them as “zero-dose”. This represents a concerning increase from 13.9 million in 2022 and a significant rise of nearly 2.7 million compared to the 12.9 million zero-dose children recorded pre-pandemic in 2019 WHO. Critically, over half of these children reside in countries affected by conflict, fragility, or instability, where access to fundamental health services is severely compromised, and the vast majority are concentrated in low– and middle-income countries, particularly within Africa and South-East Asia WHO.[8][9].

Africa faces a significant crisis in childhood immunization, leading the world in the number of children missing out on routine vaccinations. In 2021 alone, 8.7 million African children were classified as ‘zero-dose,’ having never received a single vaccine, out of a total 12.7 million under-vaccinated children on the continent.[10]. This problem is geographically concentrated, with half of the 20 countries worldwide most affected by zero-dose children situated in Africa. Within this context, Nigeria and Ethiopia, Somaliland’s neighbour, stand out, with these two nations alone home to over 3.3 million zero-dose children—representing 40% of the continent’s total [10][11]. Studies in Ethiopia, for instance, show that while full immunization coverage was low (e.g., 39% in Debie et al., 2020), significant proportions of children were also likely zero-dose or incompletely vaccinated, with spatial clustering of these vulnerable groups. [1][12][13]

Inspired by the demonstrated success of large-scale global efforts to control specific infectious diseases, the World Health Organization (WHO) launched the Expanded Program on Immunization (EPI) at the World Health Assembly in 1974[14][11][15]. The initial goal of EPI was to systematically apply and expand this successful model to immunize children against other key vaccine-preventable diseases (VPDs), thereby establishing a crucial global framework to reduce child mortality and morbidity from these illnesses [15]. The Expanded Program on Immunization (EPI) in Somaliland commenced in 1995, established in collaboration with WHO and UNICEF [16]. However, findings from the Somaliland Demographic Health Survey indicate persistent challenges, with merely 13% of children under the age of two having completed their immunization schedule [17]. Such low coverage contributes to an elevated under-five mortality rate of 117 per 1,000, a significant portion of which is attributable to vaccine-preventable diseases[18].

Identifying the geographical distribution and determinants of zero-dose status is crucial for effective public health interventions. National-level statistics often obscure significant subnational variations and inequities [11][11]. Research in similar contexts has consistently shown that zero-dose prevalence is not uniform. For example, a study in Kikwit, DRC, found a 16.3% zero-dose prevalence with significant spatial clustering and identified hotspots [11]. Similarly, analyses across 43 LMICs revealed substantial geographical disparities, with some regions in Africa and Asia exhibiting extremely low immunization rates [11]. In Kenya, distinct hotspots of zero-dose children were identified in northern arid and semi-arid lands [19], while studies in Ethiopia also highlighted geographical clustering of incomplete immunization[13][12].

Previous studies across various low– and middle-income countries, including contexts like Ethiopia, Kenya, and the DRC, indicate that the factors contributing to zero-dose status include socio-demographic aspects such as maternal education and household wealth; child-specific factors such as age; health service access indicators like distance to facilities and ANC attendance; caregiver knowledge of VPDs; and media exposure, including mobile phone usage [14][1][19][11][13][2][12]. While considerable research exists on general immunization coverage in the Horn of Africa, there is a specific need to focus on zero-dose children in Somaliland and to explore the spatial patterns and determinants associated with this status. Traditional regression analyses, while informative, may not fully capture the geographically varying nature of these determinants. Geospatial mapping, on the other hand, provides powerful tools to visualize the distribution of zero-dose children, identify high-risk areas (hotspots), and potentially uncover how determinants exert their influence differently across space.

Therefore, this study aims to utilize geospatial mapping techniques to assess the distribution and determinants of unvaccination status among children under five years in Somaliland, based on data from the 2020 Somaliland Demographic and Health Survey (SDHS). By identifying geographical hotspots and examining associated factors, this research seeks to provide actionable evidence for policymakers and public health practitioners to develop targeted strategies to reach these highly vulnerable children, thereby strengthening immunization programs and advancing health equity in Somaliland.

### Methods

#### Study design and setting

This study employed a cross-sectional design, utilizing quantitative secondary data analysis and geospatial mapping techniques to examine the distribution and identify the determinants unvaccination status among children in Somaliland. Somaliland is located in the Horn of Africa, bordered by Djibouti, Ethiopia, and Somalia, with a coastline along the Gulf of Aden. Administratively, the country is divided into six major regions (Awdal, Marodi-Jeex, Sahil, Sool, Sanaag, and Togdheer), which are further subdivided into districts.

#### Data source

The data for this study were derived from the Somaliland Health and Demographic Survey (SLHDS) 2020. This survey, the first of its kind in Somaliland, was conducted by the Central Statistics Department of the Ministry of Planning and National Development (MoP&ND) and the Ministry of Health Development (MoHD), with technical and financial assistance from the United Nations Population Fund (UNFPA) and other partners. Fieldwork for the main survey was largely undertaken between August 2018 and December 2019.

The SLHDS 2020 was designed to be nationally representative, covering all six administrative regions and stratifying by urban, rural, and nomadic populations using a multi-stage cluster sampling approach. For urban and rural areas, Enumeration Areas (EAs) were created from digitized dwelling structures via satellite imagery, followed by household listing and systematic selection. For nomadic populations, Temporary Nomadic Settlements (TNS) identified by local informants served as primary sampling units, with subsequent household listing and selection. Data were collected using standardized Household and Ever-Married Woman’s Questionnaires via Computer-Assisted Personal Interviewing (CAPI). For the present study, individual child records, their vaccination status (ascertained from vaccination cards or maternal recall), relevant socio-demographic and economic characteristics of the child, mother, and household, along with geospatial cluster coordinates (EAs/TNSs), were extracted from the SLHDS 2020 datasets.

### Variables

#### Outcome variable

The primary outcome variable for this study was the child’s vaccination status. This was determined using the SLHDS 2020 data, specifically the response to the “ever had vaccinated” question. For the purpose of this analysis, children who were reported as having received at least one dose of any routine childhood vaccine (recorded as ‘Yes’ to “ever had vaccinated” in the SDHS dataset) were classified as **Vaccinated children**. Conversely, children who were reported as having not received any routine childhood vaccines (recorded as ‘No’ to “ever had vaccinated”) were categorized as **un-vaccination children**. This binary categorization (un-vaccinated vs. vaccinated) allowed for the investigation of factors associated with children being completely missed by the vaccination program in Somaliland.

#### Predictor variables

Predictor variables were selected based on their documented influence on childhood vaccination in existing literature and their availability in the SDHS 2020 dataset. These variables were categorized into individual/household-level and community-level factors to facilitate a multilevel analysis.

At the **individual and household level**, predictors included child-specific characteristics such as the sex of the child. Maternal factors encompassed the mother’s highest educational level attained (categorized as no education, primary, secondary, or higher), her current pregnancy status (yes/no), current breastfeeding status (yes/no), the number of Antenatal Care (ANC) visits attended during her last pregnancy (grouped as no ANC visit, fewer than 4 visits, or 4 or more visits), and her exposure to media. Household characteristics considered were the place of delivery for the child (home or health facility) and the household wealth index (which was categorized into poor, middle, and rich based on the SDHS asset scores).

At the **community level**, predictors aimed to capture broader contextual influences. These included the child’s administrative region of residence (Awdal, Marodi-Jeex, Sahil, Togdheer, Sool, or Sanaag) and the type of settlement area, specifically the place of residence (categorized as urban or rural). Community education level (categorized low and high) also community ANC utilization (categorized high and low) lastly community poverty also categorized high and low.

### Data management and analysis

#### Data Source and Preparation

This study utilized secondary data from the Somaliland Health and Demographic Survey (SLHDS) 2020, a nationally representative survey employing a multi-stage cluster sampling design. Rigorous data quality assurance procedures were integral to the primary data collection. The primary outcome was a binary variable: ‘Un-vaccinated’ (children who had received no routine vaccines) versus ‘vaccinated’ (children who had received at least one). Cases with missing data on the outcome or key predictor variables were excluded from the final analysis.

#### Statistical Analysis (Determinants of Unvaccinated Status)

Since the outcome variable was Binary (yes vs. no), bivariable and multivariable multilevel binary logistic regression analyses were conducted to assess the association between zero-dose status and various predictor variables, accounting for the hierarchical structure of the DHS data. Independent variables demonstrating an association with zero-dose status at a p-value < 0.05 in the bivariable multilevel logistic regression analysis were considered for inclusion in the multivariable multilevel logistic regression model. In the final multivariable model, a 95% Confidence Interval (CI) was employed for Adjusted Odds Ratios (AORs), and a p-value < 0.05 was used to determine statistical significance.

Four multilevel logistic regression models were developed to assess determinants of zero-dose vaccination status: Model 0 (null model without predictors), Model I (individual/household-level predictors), Model II (community-level predictors), and Model III (all predictors). Model comparison and selection were guided by Akaike’s Information Criterion (AIC), Bayesian Information Criterion (BIC), and Log Likelihood values; the model demonstrating the lowest AIC, BIC, and highest Log Likelihood (or lowest deviance) was considered the best fit. Random effects, quantifying community-level variation, were evaluated using the Intra-class Correlation Coefficient (ICC) and community-level variance components from each model.

##### Spatial Autocorrelation

To explore the geographical distribution of un-vaccinated status in Somaliland, both global and local indicators of spatial correlation were employed as imperative tools.

##### Global Autocorrelations

To proceed with identifying geographical variation in zero-dose vaccination status, a global spatial autocorrelation analysis was conducted. The Global Moran’s I index was used to detect whether the observed spatial patterns of zero-dose prevalence were due to significant clustering, dispersion, or random distribution across the country. This initial exploratory spatial analysis aimed to identify the presence of geographical dependence in the distribution of un-vaccinated children in Somaliland.

##### Local Statistical Analysis

Further investigation using figures and maps was necessary, particularly if Global Moran’s I indicated a significant clustering effect (positive spatial autocorrelation) of zero-dose vaccination prevalence across Somaliland. Therefore, hotspot analysis using the Getis-Ord Gi* statistic was performed to identify specific patterns of spatial variation in zero-dose status [cite relevant reference if needed, like your ref. This analysis aimed to visually emphasize and locate areas with statistically significant high concentrations (hotspots) or low concentrations (cold spots) of zero-dose children, corroborating the overall clustering effect identified by global autocorrelation. Additionally, cluster and outlier analysis using Anselin Local Moran’s I (LISA) was employed to further describe the local spatial patterns of zero-dose vaccination prevalence. This LISA analysis served to confirm and complement the hotspot analysis by identifying significant local spatial clusters (e.g., High-High, Low-Low) and spatial outliers (High-Low, Low-High), thereby permitting a detailed identification of groupings and specific areas where significant deviations in zero-dose prevalence occur.

#### Statistical Analysis

##### As the outcome variable was binary

bivariable and multivariable multilevel logistic regression analyses were conducted to assess the association between zero-dose status and various predictor variables, accounting for the hierarchical structure of the DHS data. Independent variables demonstrating an association with unvaccinated at a p-value < 0.20 in the bivariable analysis were considered for inclusion in the final multivariable model. In the final model, a 95% Confidence Interval (CI) was employed for Adjusted Odds Ratios (AORs), and a p-value < 0.05 was used to determine statistical significance.

Four multilevel logistic regression models were developed: Model 0 (null model), Model I (individual/household-level predictors), Model II (community-level predictors), and Model III (all predictors). Model comparison was guided by Akaike’s Information Criterion (AIC) and Bayesian Information Criterion (BIC); the model demonstrating the lowest values was considered the best fit. Random effects, quantifying community-level variation, were evaluated using the Intra-class Correlation Coefficient (ICC).

#### SpatialAnalysis

To explore the geographical distribution of un-vaccination status, both global and local indicators of spatial **autocorrelation** were employed.

- **Global Spatial Autocorrelation:** The Global Moran’s I index was used to detect whether the observed spatial patterns of zero-dose prevalence were clustered, dispersed, or randomly distributed across the country.
- **Local Spatial Analysis:** To identify specific local patterns, *hotspot analysis (Getis-Ord Gi)*\* was performed to locate areas with statistically significant high concentrations (hotspots) or low concentrations (cold spots) of zero-dose children. Additionally, **cluster and outlier analysis (Anselin Local Moran’s I – LISA)** was employed to further describe local spatial patterns, identifying significant spatial clusters (e.g., High-High, Low-Low) and spatial outliers (High-Low, Low-High)

## Results

This study utilized data from a total unweighted sample of 3,255 children under five years of age. The descriptive analysis of children included in the study reveals significant variations in vaccination status across different socio-demographic and regional characteristics in Somaliland (Table 1). Overall, a substantial proportion of children (79.9%) were found to be completely unvaccinated.

**Table 1.** Distribution of Unvaccinated Status among Children Under Five by Socio-Demographic Characteristics, SLHDS 2020.

| Variable | Categories | Total N (Weighted %) | Vaccination Status |  |
| --- | --- | --- | --- | --- |
|  |  |  | Unvaccinated<br>N (%) | Vaccinated<br>N(%) |
| Regions | Awdal | 194 (5.96%) | 172 (88.66) | 22 (11.34%) |
|  | Marodijex | 213 (6.54%) | 168 (78.87%) | 45 (21.13%) |
|  | Sahil | 364 (11.18%) | 260 (71.43%) | 104 (28.57%) |
|  | Togdheer | 531 (16.81%) | 400 (75.33%) | 131 (24.67%) |
|  | Sool | 1,020 (31.34%) | 864 (84.71%) | 156 (15.29%) |
|  | Sanaag | 933 (28.66%) | 736 (78.89%) | 197 (21.11%) |
| Place of residence | Urban | 1,619 (49.74%) | 1,294 (79.93%) | 325 (20.07%) |
|  | Rural | 1,636 (50.26) | 1,306 (79.83%) | 330 (20.17%) |
| Highest Educational level | No education | 2,789 (85.68%) | 2,294 (82.25%) | 495 (17.75%) |
|  | Primary | 397 (12.20%) | 278 (70, 03%) | 119 (29.97%) |
|  | Secondary | 54 (1.66%) | 22 (40.74%) | 32 (59.26%) |
|  | Higher | 15 (0.46%) | 6 (40%) | 9 (60%) |
| Sex of child | Male | 1,723 (52.93%) | 1,394 (80.91%) | 329 (19.09%) |
|  | Female | 1,532 (47.07%) | 1,206 (78.72%) | 326 (21.28%) |
| Currently pregnant | Yes | 580 (17.82%) | 426 (73.45%) | 154 (26.55%) |
|  | No | 2,675 (82.18%) | 2,174 (81.27%) | 501 (18.73%) |
| Currently breastfeeding | Yes | 1,895 (58.22%) | 1,580 (83.38%) | 315 (16.62%) |
|  | No | 1,360 (41.78%) | 1,020 (75%) | 340 (25%) |
| Place of delivery | Home | 2,861 (82.375) | 2,258(84.22%) | 422(15.78%) |
|  | Health facility | 574 (17.63%) | 342(59.58%) | 232(40.42%) |
| Wealth index | Poor | 2,043 (62.76%) | 1,772(86.74%) | 271(13.26%) |
|  | Middle | 343 (10.54%) | 266(77.55%) | 77(22.45%) |
|  | Rich | 869 (26.70%) | 562(64.64%) | 307(35.33%) |
| Media exposure | No | 2,978 (91.94%) | 2,432(81.67%) | 546(18.33%) |
|  | Yes | 277 (8.51%) | 109(39.35%) | 168(60.65%) |
| ANC visit | No ANC visit | 2,381 (73.33%) | 2,074(87.11%) | 307(12.89%) |
|  | < 4 Visits | 633 (19.49%) | 402(63.51%) | 231(36.49%) |
|  | 4 or more visits | 233 (7.18%) | 118(50.64%) | 115(2011%) |

Regionally, Awdal (88.7%) and Sool (84.7%) exhibited the highest percentages of unvaccinated children, indicating these areas as having the most significant challenges in vaccination uptake. In contrast, the Sahil region reported the lowest proportion at 71.4%. There was minimal difference in vaccination status by place of residence, with both urban (79.9%) and rural (79.8%) areas showing similarly high proportions of unvaccinated children.

Maternal education level demonstrated a strong inverse association with vaccination status; 82.3% of children whose mothers had no education were unvaccinated, a figure that markedly decreased to 40.7% for mothers with secondary education. Similarly, children born at home had a higher prevalence of being unvaccinated (84.2%) compared to those born in a health facility (59.6%). The household wealth index was also clearly associated with vaccination status, with 86.7% of children from the poorest households being unvaccinated, compared to 64.6% in the richest households. Antenatal care (ANC) attendance showed a clear pattern: 87.1% of children whose mothers had no ANC visits were unvaccinated, decreasing to 63.5% for those with one to three visits, and further to 50.6% for children of mothers who attended four or more ANC visits.

A multilevel logistic regression analysis was conducted to examine both individual and community-level factors associated with zero-dose vaccination among children in Somaliland, as detailed in Table 2. Model 3 was selected as the final comprehensive model for interpretation because it incorporates all specified variables and demonstrates a strong statistical fit, comparable to more parsimonious alternatives, thereby offering a fuller understanding of the combined influences on zero-dose status.

**Table 2.** Multilevel logistics regression analysis of individual and community level factors associated Unvaccinated Status among Children Under Five Somaliland.

| Variables | Model 0 | Model 1 | Model 2 | Model 3 |
| --- | --- | --- | --- | --- |
|  |  | AOR (95%CI) | AOR (95%CI) | AOR (95%CI) |
| Region |  |  |  |  |
| Awdal |  |  | 1 | 1 |
| Marodijex |  |  | .4772(.2416- .9424)* | .6591 (.3572-1.2163) |
| Sahil |  |  | .3196(.1719- .5940)** | .8345 (.4688-1.485) |
| Togdheer |  |  | .3895 (.2290- .6624)** | .6663 (.4084-1.087) |
| Sool |  |  | .7083 (.4438-1.130) | 1.181 (.6976-2.001) |
| Sanaag |  |  | .4764 (.2937- .7730)** | .8111 (.5011-1.312) |
| Place of resident |  |  |  |  |
| Rural |  |  | 1 | 1 |
| Urban |  |  | .9755753 (.7087-1.342) | .8855 (.680-1.151) |
| Highest educational level |  |  |  |  |
| No education |  | 1 |  | 1 |
| Primary |  | 1.072 (7440-1.546) |  | 1.038 (.7224-1.493) |
| Secondary |  | 0423 (.2052-8729)* |  | .4093 (.1959-.85551)* |
| Higher |  | .5197 (.1377-1.960) |  | .5272 (.1384-2.007) |
| Sex of child |  |  |  |  |
| Male |  | 1 |  | 1 |
| Female |  | .9245 (7520-1.136) |  | .9224 (.7519-1.131) |
| Currently pregnant |  |  |  |  |
| Yes |  | 1 |  |  |
| No |  | 1.2594 (.9407-1.686) |  | 1.266 (.9520-1.683) |
| Currently breastfeeding |  |  |  |  |
| Yes |  | 1 |  | 1 |
| No |  | .5699 (.4521-7183)** |  | .5651 (.4507-.7084)** |
| Place of delivery |  |  |  |  |
| Home |  | 1 |  | 1 |
| Health facility |  | .6409 (.4776-.8602)** |  | .6421 (.4769-.8644)** |
| Wealth index |  |  |  |  |
| Poor |  | 1 |  | 1 |
| Middle |  | .6393(.4312--.9480) * |  | .6020 (.3959-.9154)** |
| Rich |  | .5075 (3514-7329)** |  | .4996 (.3473-.7187)** |
| Media exposure |  |  |  |  |
| No |  | 1 |  | 1 |
| Yes |  | .9532 (.6616-1.373 |  | .9938 (.6850-1.441) |
| ANC visit |  |  |  |  |
| No ANC visit |  | 1 |  | 1 |
| < 4 Visits |  | .3442 (.2605-4549)** |  | .3485 (.2611-4650)** |
| 4 or more visits |  | .2447 (.1709-.3504)** |  | .2530 (.1775- .3606** |
| Community ANC utilization |  |  |  |  |
| High Community ANC Utilization |  |  | 1 |  |
| Low Community ANC Utilization |  |  | 2.334(1.346-4.048)** | 1.543(0.893-2.666) |
| Community education level |  |  |  |  |
| High Community Education |  |  | 1 |  |
| Low Community Education |  |  | 1.00(0.365-2.735) | 0.922(0.338-2513) |
| Community poverty level |  |  |  |  |
| High poverty |  |  |  |  |
| Low poverty |  |  | 2.963(1.761-4.986)*** | 2.286(1.258-4156)** |

The analysis revealed several significant individual-level predictors. Maternal education played a key role: children whose mothers had attained a secondary level of education were approximately 59% less likely (AOR = 0.409; 95% CI: 0.1959–0.8555) to be zero-dose compared to those whose mothers had no formal education, although a similar trend for higher education did not reach statistical significance in this model. Interestingly, current breastfeeding status also emerged as significant, with children whose mothers were *not* currently breastfeeding being approximately 43.5% less likely (AOR = 0.565; 95% CI: 0.4507–0.7084) to be zero-dose than children of mothers who were currently breastfeeding. Furthermore, the place of delivery was influential; children delivered in a health facility had approximately 36% lower odds (AOR = 0.642; 95% CI: 0.4769– 0.8644) of being zero-dose compared to those born at home. Household economic status, represented by the wealth index, also demonstrated a clear protective gradient. Compared to children from poor households, those from middle-income families were about 40% less likely (AOR = 0.602; 95% CI: 0.3959–0.9154) to be zero-dose, and this protective effect was even stronger for children from rich households, who were approximately 50% less likely (AOR = 0.499; 95% CI: 0.3473–0.7187) to be zero-dose. Perhaps one of the strongest individual-level associations was observed with Antenatal Care (ANC) visits. The odds of a child being zero-dose decreased substantially with increased ANC attendance: compared to children whose mothers had no ANC visits, those whose mothers attended fewer than four visits were about 65% less likely (AOR = 0.349; 95% CI: 0.2611–0.4650) to be zero-dose, while children whose mothers had four or more ANC visits were an impressive 75% less likely (AOR = 0.253; 95% CI: 0.1775–0.3606) to be zero-dose. Conversely, factors such as the child’s place of residence (urban versus rural), the sex of the child, whether the mother was currently pregnant, and media exposure did not show statistically significant independent associations with zero-dose status in this fully adjusted model.

Beyond individual characteristics, community-level factors revealed significant contextual influences on child vaccination status. Initial analyses (Model 1) indicated notable regional disparities, with children in Marodijex, Sahil, Togdheer, and Sanaag regions demonstrating significantly lower odds of being zero-dose compared to Awdal. While a comprehensive interpretation of these regional effects within the fully adjusted Model 3 would require its specific AORs, these findings point to existing geographic variations. Regarding other community-level variables, community ANC utilization initially appeared to be a strong predictor in Model 2 (AOR = 2.334), where children in communities with low ANC utilization were more likely to be unvaccinated. However, this association’s significance diminished in the comprehensive Model 3 (AOR = 1.543), suggesting that other individual or community factors included in the final model may mediate or account for some of its initial effect. Community education level did not show a statistically significant association with zero-dose status in the fully adjusted Model 3. In stark contrast, community poverty level emerged as a robust and independent determinant; children residing in communities characterized by low poverty levels were approximately 2.29 times more likely to be unvaccinated (AOR = 2.286; 95% CI: 1.258–4.156) compared to those in high poverty communities (Model 3). This robust finding underscores that the broader socioeconomic environment of a community plays a crucial and independent role in shaping zero-dose vaccination rates in Somaliland.

**Table 3.** Model comparison and random effect analysis result.

| Parameters | Model 0 | Model I | Model II | Model III |
| --- | --- | --- | --- | --- |
| AIC | 2298.959 | 2131.629 | 2216.657 | 2120.955 |
| BIC | 2310.05 | 2209.265 | 2277.656 | 2231.863 |
| Log likelihood | -1147.4794 | -1051.8146 | -1097.3285 | -1040.4777 |
| ICC | .247833 | .0823377 | .0878475 | .0776745 |
| Variance | 1.083985 | .295185 | .3168405 | .2770595 |

Based on the model comparison statistics, four different models (Model 0, Model I, Model II, and Model III) based on several statistical parameters, including AIC, BIC, Log likelihood, ICC, and Variance. Model III appears to be the best-fitting model, as indicated by its lowest AIC (2120.955) and highest Log likelihood (–1040.4777). While Model I has a slightly lower BIC (2209.265) compared to Model III (2231.863), Model III’s advantages in AIC and Log likelihood are more substantial. Model 0, on the other hand, performs the worst across most metrics, showing the highest AIC (2298.959) and lowest Log likelihood (–1147.4794). In terms of random effects, the Intraclass Correlation Coefficient (ICC) and Variance are highest in Model 0 (.247833 and 1.083985 respectively), suggesting a greater proportion of variance explained by clustering in that model. Models I, II, and III show considerably lower ICC and Variance values, with Model III having the lowest ICC (.0776745) and Variance (.2770595), indicating less unexplained variance due to random effects compared to the other models.

**Figure 1:**
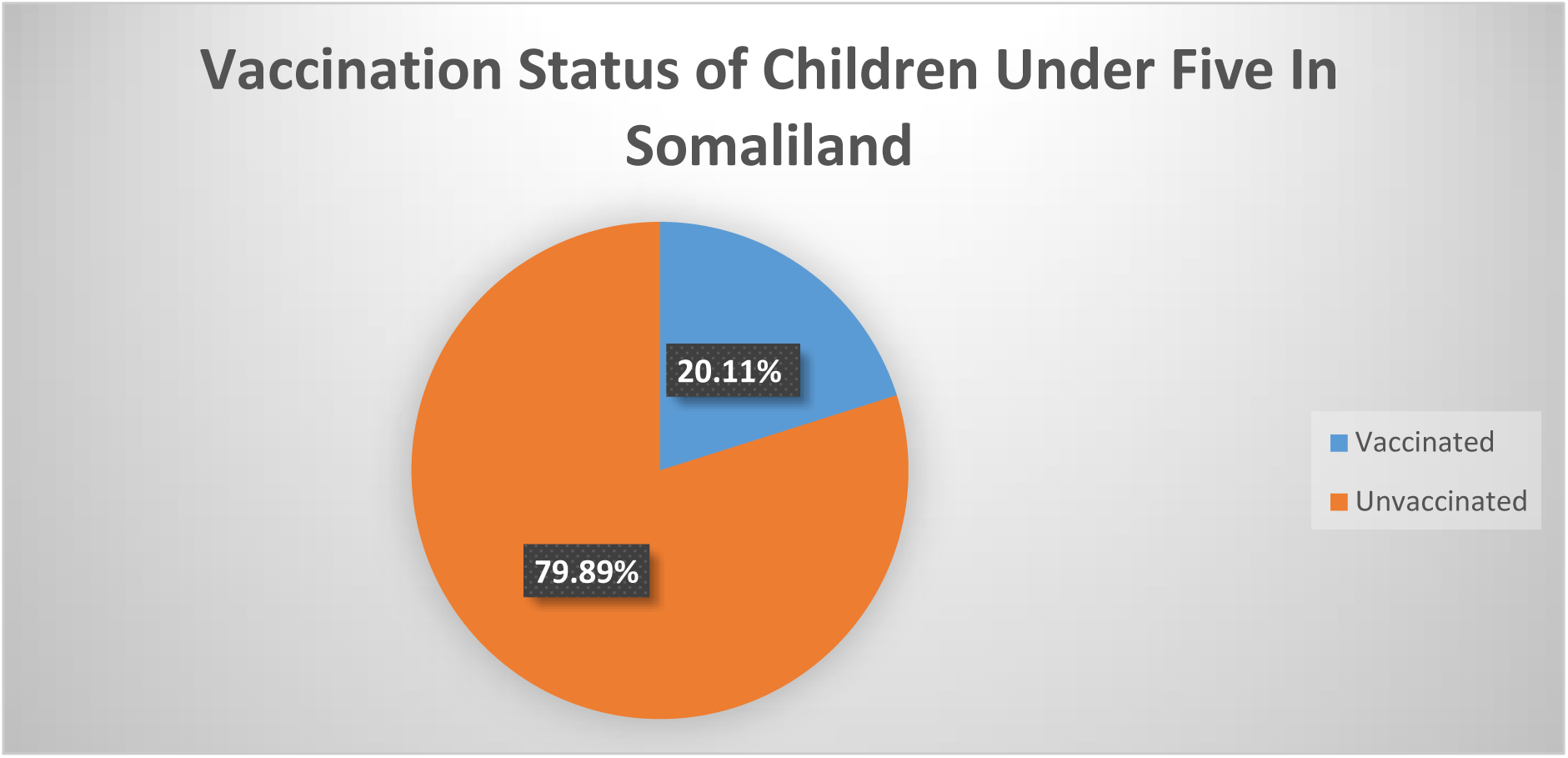
Vaccination Status of Children Under Five in Somaliland.

Figure 1 illustrates the overall vaccination status of children under five in Somaliland. The findings reveal a critical public health challenge, with a vast majority of children, 79.89%, being completely unvaccinated. This means they have not received any routine immunizations. Conversely, only a small proportion, 20.11%, are vaccinated, indicating they have received at least one vaccine dose. This highlights a substantial public health concern regarding the high number of unvaccinated children in Somaliland.

## Region in Somaliland

**Figure 2:**
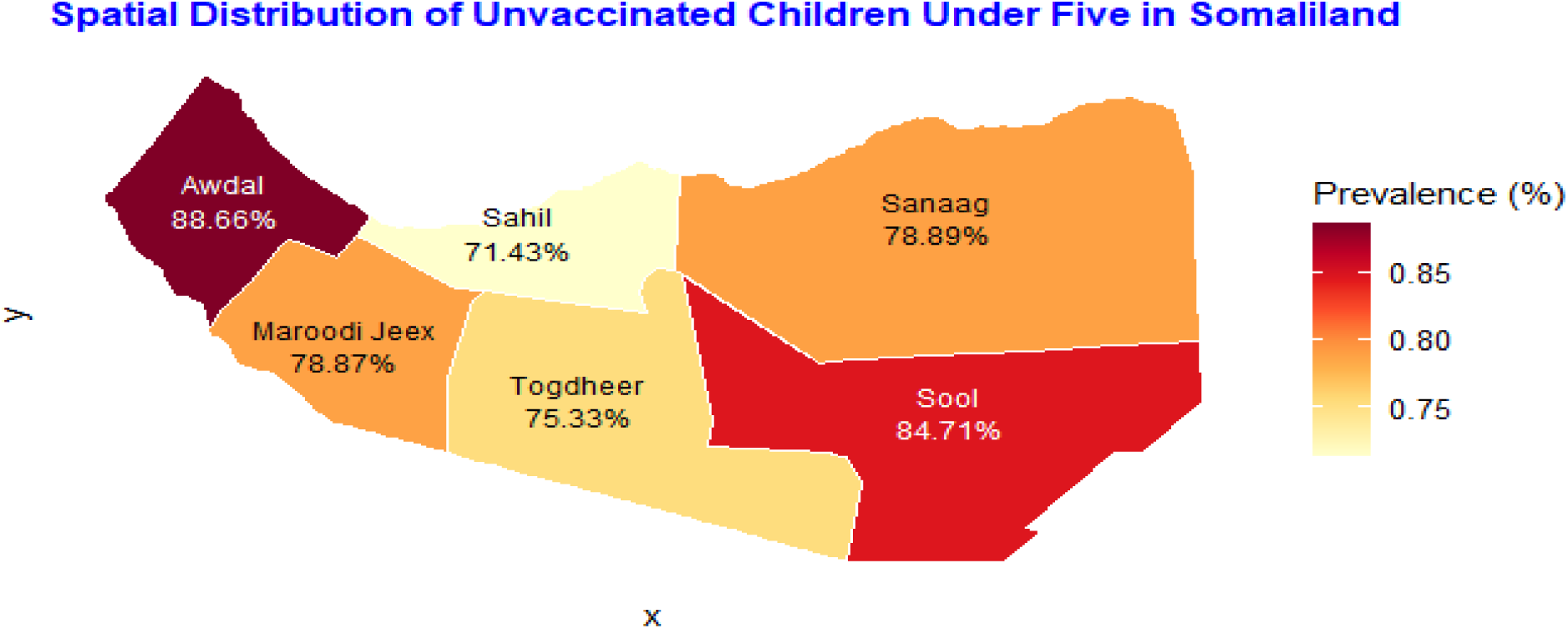
Spatial Distribution of the Prevalence of Unvaccinated Children Under Five by.

The geographical distribution of unvaccinated children under five in Somaliland, as depicted in Figure 2, reveals substantial spatial heterogeneity. The prevalence of non-vaccination is not uniform, with distinct regional hotspots of extreme vulnerability. The Awdal region in the west emerges as the most critical area, with 88.66% of its children under five being completely unvaccinated. The Sool region in the southeast also presents a severe challenge, with a prevalence of 84.71%. The eastern region of Sanaag and the central-western region of Maroodi Jeex show similarly high rates at 78.89% and 78.87%, respectively. Comparatively, the lowest rates of non-vaccination are found in the Sahil (71.43%) and Togdheer (75.33%) regions. These findings clearly demonstrate that the challenge of reaching children with routine immunizations is most acute in the westernmost and southeastern parts of Somaliland, while central areas fare relatively better.

**Figure 3:**
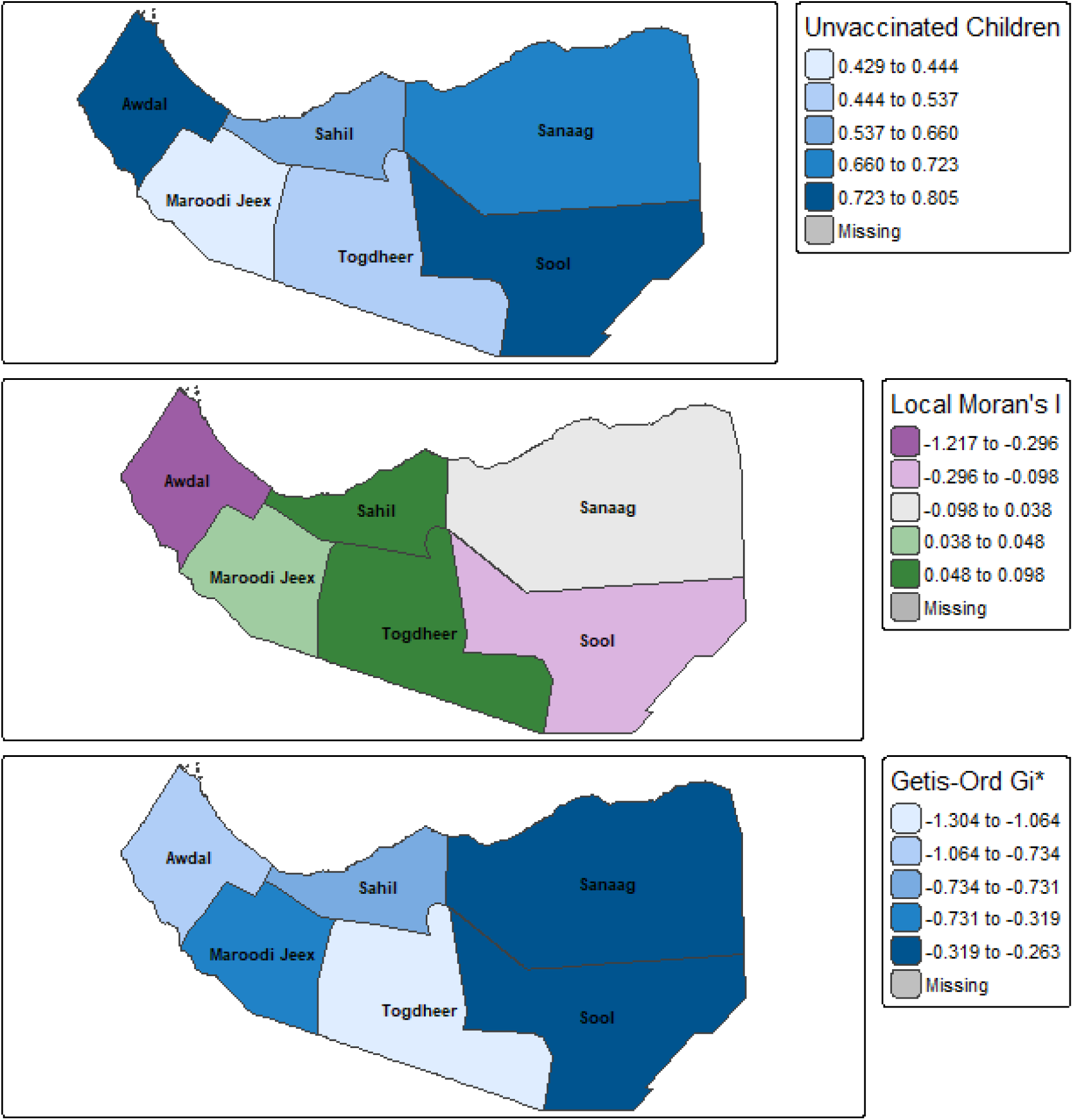
Spatial Autocorrelation Analysis of Unvaccinated Children in Somaliland. Figure 3 **presents the results of the spatial autocorrelation analysis, providing a deeper understanding of the geographic patterns of non-vaccination among children under five in Somaliland.**

The top panel displays the choropleth map of the raw prevalence of unvaccinated children, visually confirming the findings from Table 1. The darkest blue shades highlight the highest-burden regions, Sool and Sanaag, while the lightest shades indicate the regions with relatively lower prevalence, such as Togdheer and Maroodi Jeex. The middle panel illustrates the Local Moran’s I (LISA) analysis, which identifies statistically significant spatial clusters and outliers. The dark green areas, encompassing Sahil, Maroodi Jeex, and Togdheer, represent a significant Low-Low cluster. This indicates that these regions with relatively lower rates of non-vaccination are geographically clustered together. In contrast, Awdal, Sool, and Sanaag are shown in shades of purple and pink, identifying them as High-Low spatial outliers. This is a crucial finding, suggesting that these regions with a high prevalence of unvaccinated children are statistically dissimilar to their surrounding neighbors, forming “islands” of high burden rather than being part of a larger contiguous hotspot.

The bottom panel shows the Getis-Ord Gi* hotspot analysis, which identifies statistically significant spatial concentrations of high (hotspots) or low (coldspots) values. This map identifies a significant hotspot (darkest blue) in the eastern part of the country, specifically in the Sool and Sanaag regions, confirming a statistically significant concentration of high non-vaccination rates in this area. Conversely, a large coldspot (lighter blues) is identified across the western and central regions, indicating a significant spatial clustering of relatively lower non-vaccination rates, even though some individual regions within this coldspot, like Awdal, have high prevalence. Together, these maps demonstrate a complex spatial pattern defined by a significant eastern hotspot, a central-western coldspot, and notable regional outliers.

**Figure 4:**
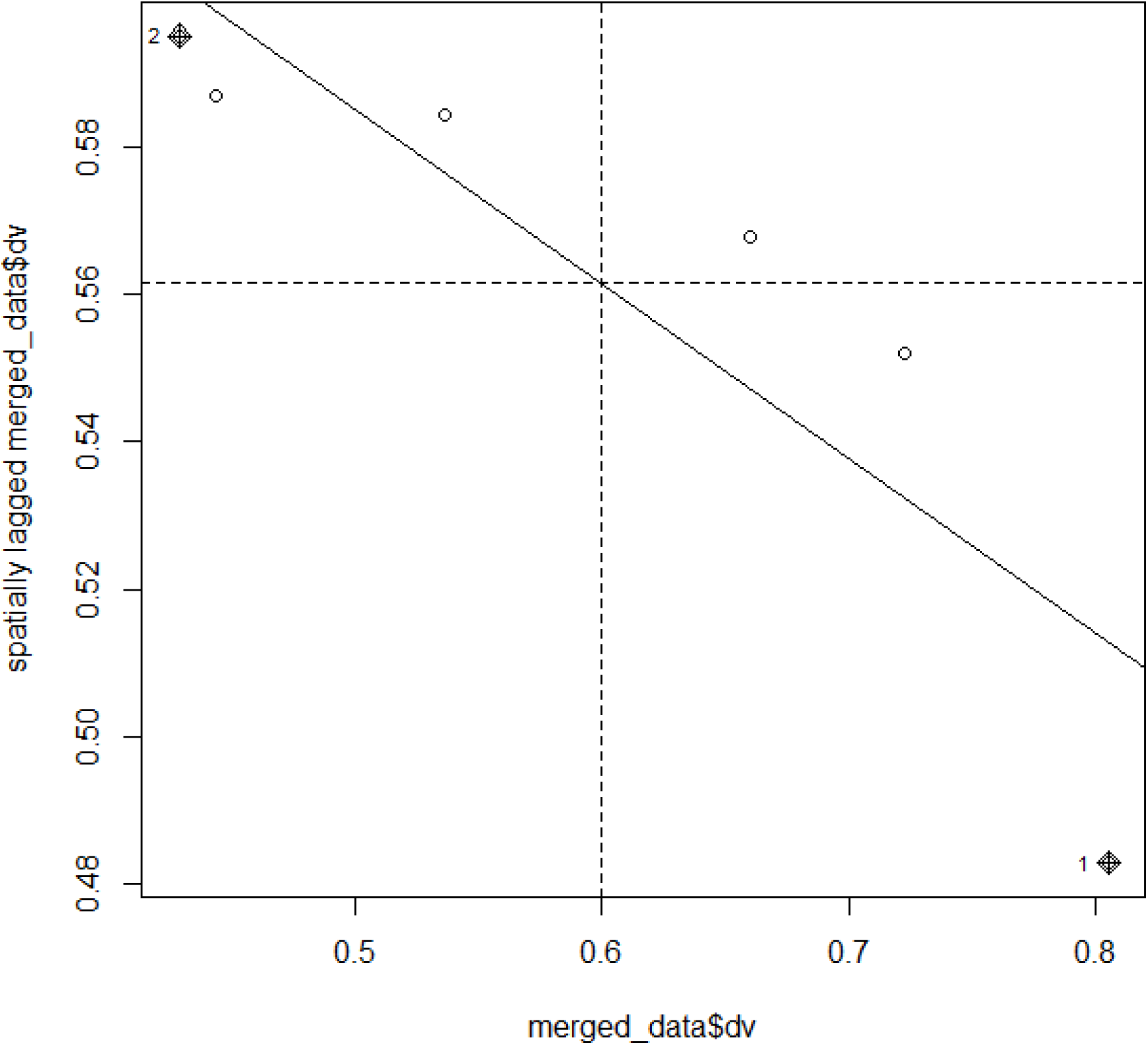
Moran Scatter Plot of the Prevalence of Unvaccinated Children in Somaliland.

This Moran scatter plot visually assesses the spatial autocorrelation of the “merged_data$dv” variable, likely representing zero-dose vaccination rates. The distinct negative slope of the overall regression line (solid diagonal) indicates a pattern of **negative global spatial autocorrelation**. This suggests that, on the whole, regions with high “dv” values tend to be located near regions with low “dv” values, and vice-versa, more frequently than would be expected by random chance, pointing towards a dissimilar or dispersed spatial pattern. Examining the local patterns within the quadrants, one region falls into the High-High quadrant, indicating a high “dv” value surrounded by similarly high-valued neighbors. However, the majority of regions display characteristics of spatial outliers: three regions, including one labeled “2”, are in the Low-High quadrant (low “dv” value surrounded by high-value neighbors), and two regions, including one labeled “1”, are in the High-Low quadrant (high “dv” value surrounded by low-value neighbors). The absence of any regions in the Low-Low quadrant further underscores the lack of clustering of low “dv” values. Therefore, while one instance of positive local clustering exists, the predominant spatial structure is characterized by dissimilarity, with most regions being outliers relative to their surrounding areas.

## Discussion

This study provides a critical spatial and multilevel analysis of un-vaccination status in Somaliland, revealing an alarmingly high prevalence of 79.88% among under five children. This figure establishes Somaliland as a profound global outlier, dwarfing the 16.5% zero-dose prevalence found across 33 Sub-Saharan African countries and the pooled 7.7% across 92 LMICs [20, 11]. The situation appears far more severe than in neighboring countries like Ethiopia, where studies have documented significant challenges with incomplete and untimely vaccination but have not reported zero-dose rates of this magnitude [28, 29, 6]. While countries like Togo have shown slow but steady progress in reducing their zero-dose burden over decades [30], the sheer scale of the problem in Somaliland underscores the critical importance of the government’s commitment to “scale up priority interventions such as the vaccination program” [21].

Our multilevel analysis identified powerful determinants that align with established evidence. The protective effect of maternal education is a consistent theme across the region, validated by research in Ethiopia, Kenya, and Nigeria [27, 19, 5]. and by Somaliland’s National Health Policy, which identifies “High illiteracy” as a key social determinant of health[22]. Similarly, contact with the health system via Antenatal Care (ANC) and facility-based delivery emerged as a highly significant protective factor. This finding is particularly poignant in light of global evidence that nearly half of children defined as “penta-zero dose” have had at least one other vaccine contact, framing ANC and delivery as critical but often missed opportunities for immunization [23, 26].

Socioeconomic status was another critical determinant, reflecting deep-rooted inequities central to Somaliland’s UHC goals [24]. This wealth-based disparity is a pervasive driver of under-vaccination across Africa [20, 6]. Importantly, this inequity has consequences that extend far beyond immediate health. Research from India demonstrates that being a zero-dose child is a marker of systemic disadvantage that predicts poorer long-term learning attainment in preadolescence, with effects most pronounced in the poorest communities [25]. This reframes Somaliland’s zero-dose crisis as a fundamental threat to its future human capital.

A key contribution of this study is the granular geospatial analysis, which revealed significant heterogeneity and identified Awdal and Sool as high-burden regions. The use of spatial modeling to identify such hotspots is a critical strategy now widely employed across Africa, with studies from Kenya[19], Zambia [26], Nigeria [27, 31], and Ethiopia [28] all demonstrating its utility for programmatic targeting. The finding of a negative global Moran’s I, suggesting a uniquely fragmented landscape, likely reflects the systemic realities of a health system reliant on a patchwork of partners, as described in the UHC Roadmap [24].

The profound influence of community context was quantified by our random-effects analysis (ICC=24.8%). This finding is powerfully illustrated by evidence that the negative developmental impacts of being zero-dose are moderated by community context, demonstrating that a child’s environment shapes their life-course outcomes [25]. The significant unexplained community-level variance in our model confirms that unobserved characteristics—such as the systemic weaknesses outlined in Somaliland’s policy documents—play a crucial role, reinforcing the need for community-based approaches, a conclusion also reached in multilevel analyses in Nigeria and across SSA [27, 20, 13].

This study’s strengths lie in its use of a recent, nationally representative dataset and robust analytical methods. Limitations must be acknowledged, including the cross-sectional design and reliance on maternal recall, a practice necessitated by the “low card retention rate” officially noted in Somaliland’s EPI policy [21]. Furthermore, like many surveys, we define zero-dose by non-receipt of DTP1, which may not perfectly distinguish the “truly zero-dose” from those with minimal health system contact [23].

In conclusion, this study provides an evidence base to operationalize the very policies designed to address Somaliland’s zero-dose crisis. Our findings strongly support a dual strategy that is not only data-driven but also directly aligned with national health priorities. First, broad initiatives to enhance female education and promote ANC utilization directly support the goals outlined in the National Health Policy III and UHC Roadmap. Second, a data-driven, geographically targeted approach for Awdal and Sool executes the precise strategies proven effective in other high-burden contexts[26,19]. By leveraging these spatial and multilevel insights, Somaliland can turn the ambitious goals of its national policies into life-saving—and life-altering—realities for its most vulnerable children, a critical step toward strengthening its health system and aligning with the global Immunization Agenda 2030[25].

## Policy Implications

The alarmingly high prevalence of unvaccinated children in Somaliland, particularly in regions like Awdal and Sool, necessitates targeted and geographically specific interventions. Public health programs should prioritize these high-burden areas with tailored strategies, such as mobile vaccination clinics, community outreach, and localized communication campaigns, to overcome specific barriers to immunization. This approach aligns with successful strategies implemented in other high-burden contexts.

Strengthening maternal education and enhancing healthcare utilization are crucial policy areas. The strong association between maternal secondary education, antenatal care (ANC) visits, and facility-based delivery with a reduced likelihood of zero-dose status underscores the need for policies that improve female education access and retention. Furthermore, integrating immunization information and services into ANC and delivery care can leverage these existing health contacts as vital opportunities for vaccination, ensuring that mothers receive essential information and children are vaccinated at birth or soon after.

Addressing socioeconomic disparities is also paramount, as household wealth is a significant determinant, with children from poorer households being more prone to being unvaccinated. Policies should tackle broader socioeconomic inequities through social protection programs, subsidies for healthcare access, and initiatives that reduce financial barriers to immunization. Recognizing that zero-dose status is a marker of systemic disadvantage, interventions should aim to improve long-term outcomes for these vulnerable populations. The substantial community-level influence on zero-dose status also highlights the importance of community-based approaches. Policies should support community health workers and local leaders in promoting immunization, addressing misinformation, and facilitating access to vaccination services, ensuring that local community dynamics are understood and addressed for effective program implementation.

Finally, the utility of geospatial analysis in identifying hotspots and cold spots provides a powerful tool for public health planning. Policymakers should integrate such data-driven approaches into national health planning to enable continuous monitoring of immunization coverage and rapid response to emerging inequities. This supports Somaliland’s National Health Policy and UHC Roadmap goals, which identify “High illiteracy” as a key social determinant of health and emphasize the need for data-driven strategies.

## Conclusion

Somaliland faces an exceptionally high prevalence of zero-dose children, with nearly 80% of children under five years of age having received no routine immunizations. This alarming figure significantly surpasses regional and global averages, establishing Somaliland as a profound global outlier in immunization coverage. The study highlights significant spatial heterogeneity in vaccination status, with regions like Awdal and Sool demonstrating particularly high rates of unvaccinated children. Key individual-level determinants include maternal education, household wealth, antenatal care attendance, and delivery in a health facility, all of which are protective factors against zero-dose status. Community-level factors also play a crucial role, underscoring the need for comprehensive and context-specific interventions. These findings provide a robust evidence base for policymakers to implement data-driven and geographically targeted strategies, focusing on enhancing maternal education, improving access to and utilization of healthcare services, and addressing socioeconomic disparities, particularly in high-burden regions. By leveraging these insights, Somaliland can strengthen its immunization programs and progress towards achieving health equity for its most vulnerable children, aligning with global immunization agendas.

## Data Availability

Data are available from department of central statistics in ministry of planning and development in Hargeisa Somaliland

